# Methamphetamine-associated Heart Failure: Clinical phenotypes and Outcomes in a Safety Net Population

**DOI:** 10.1101/2024.04.22.24306200

**Authors:** Anjali B. Thakkar, Matthew S. Durstenfeld, Yifei Ma, Sithu Win, Priscilla Y. Hsue

## Abstract

**Background:** Methamphetamine use has increased dramatically over the past decade and is associated with the development of heart failure (HF). However, clinical characteristics and outcomes have not been well described. This study aimed to compare clinical characteristics and outcomes among individuals with HF who do and do not use methamphetamines in a safety-net hospital.

**Method:** This retrospective matched cohort study included all individuals with HF with history of methamphetamine use and age, gender-, and year-matched controls without history of methamphetamine use within a municipal health system from 2001-2019. 1,783 individuals with methamphetamine use and HF were identified; 12 were excluded due to inability to identify matched methamphetamine-negative controls. Therefore, 1,771 individuals with methamphetamine use and heart failure and 3,542 age, sex, and year-of-HF-diagnosis matched controls with heart failure without methamphetamine use were included. The primary outcome was all-cause mortality. Secondary outcomes included time to HF hospitalization, and 30-day, 90-day, and 1 year HF and all-cause readmissions.

**Results:** Median age of the cohort was 52.1 years and 22.6% were female. There was no significant difference in mortality between the two groups (40% vs 36.6%, HR 1.00, 95% CI 0.91, 1.10, p=1.00). A subset had an index HF hospitalization (n=1,404) during the study period including 637 (35.9%) with history of methamphetamine use and 767 (21.7%) without history of methamphetamine use (relative risk 1.66, 95%CI 1.52-1.81, p<0.0001). Among those ever hospitalized for HF, individuals with methamphetamine use had increased odds of HF and all-cause readmission at 30 days, 90 days, and 1 year.

**Conclusion:** Despite having higher risk of both all cause and HF readmissions, individuals with methamphetamine-associated heart failure did not have higher risk of mortality. Measures to address frequent healthcare utilization among people with methamphetamine use and HF are needed.

## Introduction

Methamphetamine is a highly-addictive central nervous system stimulant which has become a global epidemic, with particularly high usage in the western United States. The Center for Disease Control reports that overdose deaths involving methamphetamine use have increased by 29% annually over the past decade.^1^ Aside from mortality, amphetamine-related hospitalizations and costs have significantly increased over the past three decades.^2^ The association between methamphetamine use and cardiomyopathy is particularly important considering the high mortality and resource utilization due to HF.^3^

Individuals with methamphetamine use are at increased risk of developing heart failure due to the deleterious effects of methamphetamine on cardiac myocytes.^4^ Chronic methamphetamine use has previously been associated with malignant hypertension, increased risk of myocardial infarction and stroke, cardiac arrhythmia, and heart failure.^4^ However, the long term impact of methamphetamine use on heart failure outcomes has not been described.

Our study aims to elucidate the impact of methamphetamine use on heart failure outcomes and mortality in a large cohort of individuals within an urban, safety-net hospital system.

## Methods

### Study design

We conducted a retrospective matched cohort study between 1/1/2001-8/1/2019 of all individuals with heart failure and a history of methamphetamine use, and age and gender-matched controls with heart failure without a history of methamphetamine use. All participants were recruited from a single municipal health system (San Francisco Health Network) in San Francisco, California that includes an academic hospital and a large number of community clinics. Data for this cohort were extracted with the assistance of the UCSF Clinical Translational Science Institute. The University of California San Francisco institutional review board approved this study.

### Participants

We created a registry of individuals with and without methamphetamine use with the goal of comparing heart failure outcomes between these populations, and determining comorbidities associated with outcomes. A diagnosis of prevalent heart failure was defined as presence of HF by ICD 9 or 10 code (ICD 9: 428, 428.0, 428.1, 428.2X, 428.3X, 428.4, 428.9, 402.01, 402.11, 402.91, 404.01, 404.03, 404.11, 404.13, 404.91, 404.93; ICD 10 I50.1, I50.20, I50.21, I50.22, I50.23, I50.30, I50.31, I50.32, I50.33, I50.40, I50.41, I50.42, I50.43, I50.9, I11.0) during either an outpatient or inpatient healthcare encounter. Methamphetamine-use status was defined as an ICD 9 (304.40-304.43; 305.70-305.73; 305.75) or ICD 10 (F15.10, F15.20) diagnosis code for methamphetamine use, or the presence of an amphetamine derivative on a urine toxicology screen. Duration of methamphetamine use could not be obtained from available data. All individuals with methamphetamine use and HF during the specified study period were included in this study, and age and gender-matched controls without history of methamphetamine use were identified. A random subset of the cohort was adjudicated by a single physician (ABT) to improve EHR-based data queries for alignment with manual chart review.

Individuals with HF and methamphetamine use were matched 1:2 with individuals with HF without methamphetamine use based on age, gender, and the date of heart failure diagnosis within a 2-year window.

### Outcomes

The primary outcome was all-cause mortality following a diagnosis of heart failure. Participants were followed from the date of HF diagnosis through the end of the study period (8/1/2019) or until date of death, whichever came first. Vital status and date of death were ascertained using both the National Death Index (NDI) and the Social Security Death Index (SSDI), and hospital death records, with matches for 3,691 (70%) participants. If no mortality data was available, the subject was censored at date of last contact. If mortality data was available from both sources with discrepant information, NDI/SSDI data was used.

Secondary outcomes included proportion with an index heart failure hospitalization, time to index HF hospitalization, and 30-day, 90-day, and 1 year heart failure and all-cause readmissions. Index HF hospitalization was defined as the patient’s first hospitalization for a primary diagnosis of HF (based on ICD 9/10 codes) during or after the date of HF diagnosis. Heart failure readmissions were defined as hospitalizations with a primary diagnosis of heart failure, based on ICD 9/10 codes, after the discharge date of the index HF hospitalization. All-cause readmissions were defined as all hospitalizations after the discharge date of the index HF hospitalization, inclusive of hospitalizations with a primary diagnosis of heart failure.

### Covariates

All covariates were obtained by ICD 9/10 codes. Cardiovascular risk factors included hypertension, hyperlipidemia, diabetes, body mass index, and obstructive sleep apnea. Cardiovascular disease included coronary artery disease, prior stroke/cerebrovascular accident, pulmonary hypertension, history of myocardial infarction, history of percutaneous coronary intervention, history of coronary artery bypass graft, and presence of arrhythmias (atrial fibrillation, atrial flutter, sustained ventricular tachycardia, ventricular fibrillation). Health habits included use of tobacco, alcohol, opiates, cocaine, cannabis, hallucinogens, sedatives/anxiolytics, or other drugs not otherwise specified. Other comorbid conditions included co-infection with hepatitis C, HIV, chronic kidney disease, chronic pulmonary disease, and depression. Comorbidities were queried from the beginning of the study period up to one year following the date of heart failure diagnosis.

HF type was classified as HFrEF or HFpEF based on estimated values of left ventricular ejection fraction extracted from clinical echocardiogram reports, with >=40% classified as HFpEF and <40% classified as HFrEF. When multiple echocardiograms were available, data was obtained from the echocardiogram closest to the date of HF diagnosis. Data on prescription of goal-directed heart failure therapies were also abstracted from the medical record.

### Statistical analysis

We first compared demographic and clinical characteristics of individuals with and without methamphetamine use for the entire sample and the subset with index hospitalization. For categorical variables, counts and percentages were calculated and chi-square test was performed. For continuous variables, Shapiro-Wilk normality test was performed, and either t-test or Wilcoxon rank test was performed where appropriate. Key lab characteristics, medication use and hospitalization characteristics were also compared among individuals with index hospitalization. Heart failure and all cause hospital readmission rate at 30-day, 90-day, and 1-year was also estimated and relative risks of readmission were compared between individuals with and without methamphetamine use among those with index hospitalization. Methamphetamine-use stratified cumulative incidence of all-cause readmissions, HF readmissions, and mortality of individuals were presented using Kaplan Meier curves. Cox regression models were used to estimate the hazard ratio of methamphetamine use on mortality. Fine-Gray competing risk models were used to provide estimation of hazard ratios for the hospital readmissions over time as death presents a competing risk. The main predictors include use of methamphetamine, demographic factors, insurance status, clinical factors, comorbidities, tobacco and alcohol use, and presence of 30-day outpatient follow-up. A stepwise backward selection approach was used to consider adjusting for other comorbid conditions; that model included age, race, gender, coronary artery disease, hepatitis B virus, chronic kidney disease, and history of sustained ventricular tachycardia **(Supplementary Table 1).** Interaction terms between methamphetamine use and key covariates of interest were examined and included if significant. Subgroup analysis based on HF type was also performed. All the analyses used two-sided tests and p<0.05 was considered to be statistically significant. We used SAS 9.4 (SAS Institute, Cary, NC) software for all the analyses.

## Results

### Demographic and clinical characteristics

From 2001 to 2019, we identified 1,783 individuals with methamphetamine use and HF; 12 were excluded due to inability to identify matched methamphetamine-negative controls based on age, gender, and date of index HF hospitalization. We therefore included 1,771 individuals with methamphetamine use and heart failure, and 3,542 age, sex, and year-of-HF-diagnosis matched controls with heart failure without methamphetamine use. A subset of the cohort had an index heart failure hospitalization (n=1,404) including 637 (35.9%) with methamphetamine use and 767 (21.7%) individuals without history of methamphetamine use (relative risk [RR]=1.66, 95%CI 1.52-1.81, p<0.0001).

Median age was 52.1 years and 22.6% were female. Individuals with methamphetamine use were more likely to be Black/African American, English speaking, and insured by Medicaid **(Table 1).** Methamphetamine use was associated with both cardiovascular and non-cardiovascular comorbidities including hypertension, pulmonary hypertension, vascular disease, hepatitis C, hepatitis B, and HIV. Methamphetamine use was also associated with other substance use including alcohol, opiates, and cocaine. A higher proportion of individuals with methamphetamine use also had documented follow up within 30 days (62.4% vs 56.7%; RR 1.10, 95%CI 1.05 to 1.10, p<0.0001). Demographic and clinical characteristics among individuals with an index HF hospitalization were similar to the larger cohort (**Table 1).**

**Table 1.** Baseline characteristics of participants included in study.

|  | Methamphetamine Use |  | No Methamphetamine Use |  | P-value |  |
| --- | --- | --- | --- | --- | --- | --- |
|  | Total<br>(n=1,771) | Ever<br>hospitalized<br>(n=637) | Total (n=3542) | Ever<br>hospitalized<br>(n=767) | Total | Ever<br>hospitalized |
| **Demographic** |  |  |  |  |  |  |
| Mean age, years (SD)* | 52.1 (9.9) | 51.0 (9.5) | 52.1 (9.9) | 51.1 (9.8) | 0.980 | 0.901 |
| Race |  |  |  |  | <.0001 | <.0001 |
| White, % | 524 (29.6%) | 132 (20.7%) | 944 (26.7%) | 152 (19.8%) |  |  |
| Black/African American, % | 870 (49.1%) | 322 (50.5%) | 1169 (33%) | 301 (39.2%) |  |  |
| Asian, % | 174 (9.8%) | 110 (17.3%) | 651 (18.4%) | 146 (19%) |  |  |
| Latino/Other**, % | 203 (11.4%) | 73 (11.5%) | 778 (22%) | 168 (21.9%) |  |  |
| Gender* |  |  |  |  | 1.000 | 0.089 |
| Female, % | 401 (22.6%) | 110 (17.3%) | 802 (22.6%) | 160 (20.9%) |  |  |
| Male, % | 1370 (77.4%) | 527 (82.7%) | 2740 (77.4%) | 607 (79.1%) |  |  |
| Language |  |  |  |  | <.0001 | <.0001 |
| Non-English, % | 71 (4%) | 28 (4.4%) | 646 (18.2%) | 119 (15.5%) |  |  |
| English % | 1700 (96%) | 609 (95.6%) | 2896 (81.8%) | 648 (84.5%) |  |  |
| Insurance Type |  |  |  |  | <.0001 | <.0001 |
| Medicare, % | 527 (29.8%) | 176 (27.6%) | 958 (27%) | 211 (27.5%) |  |  |
| Medicaid, % | 956 (54%) | 378 (59.3%) | 1490 (42.1%) | 364 (47.5%) |  |  |
| Commercial, % | 6 (0.3%) | 1 (0.2%) | 74 (2.1%) | 11 (1.4%) |  |  |
| Uninsured, % | 73 (4.1%) | 28 (4.4%) | 222 (6.3%) | 56 (7.3%) |  |  |
| Unknown | 209 (11.8%) | 54 (8.5%) | 798 (22.5%) | 125 (16.3%) |  |  |
| Smoker, % | 1433 (80.9%) | 551 (86.5%) | 1613 (45.5%) | 476 (62.1%) | <.0001 | <.0001 |
| Ejection fraction |  |  |  |  | <.0001 | <.0001 |
| HFpEF (EF>40%) | 821 (54.0%) | 149 (24.5%) | 1639 (61.6%) | 230 (32.7%) |  |  |
| HFrEF (EF≤40%) | 698 (46.0%) | 459 (75.5%) | 1020 (38.4%) | 474 (67.3%) |  |  |
| **Cardiovascular comorbidities** |  |  |  |  |  |  |
| Hypertension, % | 1452 (82%) | 152 (23.9%) | 2750 (77.6%) | 186 (24.3%) | 0.000 | 0.865 |
| CAD, % | 395 (22.3%) | 79 (12.4%) | 829 (23.4%) | 91 (11.9%) | 0.368 | 0.759 |
| Pulmonary Hypertension, % | 197 (11.1%) | 107 (16.8%) | 274 (7.7%) | 118 (15.4%) | <.0001 | 0.472 |
| History of MI, % | 277 (15.6%) | 34 (5.3%) | 510 (14.4%) | 56 (7.3%) | 0.230 | 0.135 |
| History of PCI, % | 108 (6.1%) | 4 (0.6%) | 289 (8.2%) | 7 (0.9%) | 0.008 | 0.547 |
| History of CABG, % | 14 (0.8%) | 51 (8%) | 51 (1.4%) | 70 (9.1%) | 0.042 | 0.457 |
| DM, % | 169 (9.5%) | 11 (1.7%) | 393 (11.1%) | 9 (1.2%) | 0.083 | 0.384 |
| Cerebrovascular disease | 46 (2.6%) | 33 (5.2%) | 72 (2%) | 31 (4%) | 0.188 | 0.308 |
| Vascular disease | 124 (7%) |  | 185 (5.2%) |  | 0.009 |  |
| **Other Comorbidities** |  |  |  |  |  |  |
| Hepatitis C, % | 793 (44.8%) | 33 (5.2%) | 484 (13.7%) | 27 (3.5%) | <.0001 | 0.126 |
| Hepatitis B, % | 151 (8.5%) | 67 (10.5%) | 140 (4%) | 112 (14.6%) | <.0001 | 0.022 |
| CKD, % | 241 (13.6%) | 38 (6%) | 535 (15.1%) | 58 (7.6%) | 0.145 | 0.238 |
| Obstructive Sleep Apnea, % | 67 (3.8%) | 11 (1.7%) | 172 (4.9%) | 9 (1.2%) | 0.075 | 0.384 |
| Prior stroke/cerebrovascular disease, % | 46 (2.6%) | 5 (0.8%) | 72 (2%) | 7 (0.9%) | 0.188 | 0.796 |
| COPD, % | 17 (1%) | 66 (10.4%) | 20 (0.6%) | 33 (4.3%) | 0.102 | <.0001 |
| HIV, % | 257 (14.5%) | 551 (86.5%) | 165 (4.7%) | 476 (62.1%) | <.0001 | <.0001 |
| Cannabis use, % | 332 (18.7%) | 120 (18.8%) | 222 (6.3%) | 74 (9.6%) | <.0001 | <.0001 |
| Alcohol use disorder, % | 928 (52.4%) | 331 (52%) | 859 (24.3%) | 238 (31%) | <.0001 | <.0001 |
| Opioid use disorder, % | 771 (43.5%) | 254 (39.9%) | 352 (9.9%) | 84 (11%) | <.0001 | <.0001 |
| Cocaine use, % | 1028 (58%) | 394 (61.9%) | 521 (14.7%) | 192 (25%) | <.0001 | <.0001 |
| Hallucinogen use, % | 32 (1.8%) | 10 (1.6%) | 2 (0.1%) | 2 (0.3%) | <.0001 | 0.008 |
| Sedative use, % | 80 (4.5%) | 17 (2.7%) | 22 (0.6%) | 5 (0.7%) | <.0001 | 0.002 |
| Other drug use, % | 1212 (68.4%) | 431 (67.7%) | 579 (16.3%) | 164 (21.4%) | <.0001 | <.0001 |
| **Arrhythmia History** |  |  |  |  |  |  |
| Atrial fibrillation, % | 256 (14.5%) | 106 (16.6%) | 688 (19.4%) | 145 (18.9%) | <.0001 | 0.270 |
| Atrial flutter, % | 75 (4.2%) | 35 (5.5%) | 141 (4%) | 41 (5.3%) | 0.658 | 0.902 |
| Sustained ventricular tachycardia, % | 45 (2.5%) | 22 (3.5%) | 117 (3.3%) | 29 (3.8%) | 0.128 | 0.744 |
| Ventricular fibrillation, % | 13 (0.7%) | 6 (0.9%) | 58 (1.6%) | 7 (0.9%) | 0.007 | 0.955 |
| Presence of 30 day outpatient follow up | 1105 (62.4%) | 416 (65.3%) | 2009 (56.7%) | 529 (69%) | <.0001 | 0.145 |

*\*The methamphetamine use and no methamphetamine use cohorts were matched by age, gender, and year of heart failure diagnosis.*
*\*\*Latino & Other race categories are reported together as nearly all individuals with race categorized as “Other” were inferred to be Hispanic/Latino due to primary language being Spanish.*
*HFpEF = Heart failure with preserved ejection fraction; HFrEF = Heart failure with reduced ejection fraction; CAD = coronary artery disease; MI = myocardial infarction; PCI = percutaneous coronary intervention; CABG = coronary artery bypass graft; DM = diabetes mellitus; CKD = chronic kidney disease; COPD = chronic obstructive pulmonary disease; HIV = human immunodeficiency virus.*

### Heart failure characteristics

HFpEF was more prevalent than HFrEF in both individuals with (n=821; 54% vs n=698; 46%, p<0.001) and without (n=1639; 61.6% vs n=1020; 38.4%, p<0.001) methamphetamine use as demonstrated in **Table 1**. Baseline heart failure characteristics of individuals with an index heart failure hospitalization are summarized in **Tables 2a & 2b.** Key baseline heart failure lab (sodium, troponin, brain natriuretic peptide) values were similar between groups for both HFrEF and HFpEF. Among the 933 individuals with HFrEF in whom heart failure data were available, a greater percentage of individuals with history of methamphetamine use had an ambulatory prescription for guideline directed medical therapy (GDMT) for HFrEF—specifically beta blocker, ACE inhibitor/ARB/ARNi, or MRA—compared to individuals without history of methamphetamine use (44.6% vs 23.6%; RR 1.89; 95% CI 1.64 to 2.16, p<0.0001). Among individuals with CAD, prior PCI, or prior CABG, individuals with history of methamphetamine use had greater percentage of being prescribed aspirin, statin, and beta blocker compared to individuals without history of methamphetamine use regardless of HF type (HFrEF vs HFpEF). *Mortality by methamphetamine use status*

Among the entire cohort, there was no significant difference in mortality between individuals with and without methamphetamine use over a median follow up of 31.6 months (IQR: 9.5 to 69.6 months) as demonstrated in **Figure 1a** (40%; n=698 vs 36.6%; n=1167, HR 1.00, 95% CI 0.91, 1.10, p=1.00). There was also no significant difference in mortality by methamphetamine use status among those with an index HF hospitalization (45.8%; n=285 vs 44.6%; n=318, HR 0.92, 95% CI 0.78 to 1.08, p=0.29; **Figure 1b).**

**Table 2a.** Heart failure characteristics of subgroup of study participants with HFrEF.

|  | **Meth+ (n=459)** | **Meth- (n=474)** | **P value** |
| --- | --- | --- | --- |
| **Key lab characteristics** | **Median (IQR)** | **Median (IQR)** |  |
| Serum sodium | 140 (138, 142) | 141 (138, 142) | 0.049 |
| Serum troponin | 0.1 (0.1, 0.2) | 0.1 (0.1, 0.2) | 0.283 |
| Serum BNP | 1126.5 (252, 1725) | 535 (156, 1090) | 0.195 |
| **Medication Use During Study Period** | **N (%)** | **N (%)** |  |
| Aspirin, % | 216 (47.1%) | 165 (34.8%) | 0.0001 |
| Statin, % | 206 (44.9%) | 163 (34.4%) | 0.0001 |
| Beta blocker % | 289 (63%) | 224 (47.3%) | <.0001 |
| ACE inhibitor % | 282 (61.4%) | 205 (43.2%) | <.0001 |
| ARB % | 51 (11.1%) | 56 (11.8%) | 0.736 |
| ARNI % | 16 (3.5%) | 14 (3%) | 0.645 |
| Loop diuretic % | 292 (63.6%) | 235 (49.6%) | <.0001 |
| Mineralocorticoid receptor antagonist % | 150 (32.7%) | 118 (24.9%) | 0.009 |
| Hydralazine % | 51 (11.1%) | 45 (9.5%) | 0.461 |
| Imdur % | 55 (12%) | 52 (11%) | 0.628 |
| Digoxin % | 20 (4.4%) | 13 (2.7%) | 0.182 |

*\* Laboratory data was limited by availability in the hospital medical records and values were not collected for all participants. Data for creatinine and potassium were unreliable on manual adjudication and therefore were not reported.*

**Table 2b.** Heart failure characteristics of subgroup of study participants with HFpEF.

|  | **Meth+ (n=149)** | **Meth- (n=230)** | **P value** |
| --- | --- | --- | --- |
| **Key lab characteristics** | **Median (IQR)** | **Median (IQR)** |  |
| Serum sodium | 140 (138, 142) | 141 (139, 143) | 0.024 |
| Serum troponin | 0.1 (0.1, 0.1) | 0.1 (0.1, 0.2) | 0.799 |
| Serum BNP | 238.5 (34, 727) | 581 (109, 842) | 0.613 |
| **Medication Use During Study Period** | **N (%)** | **N (%)** |  |
| Aspirin, % | 66 (44.3%) | 103 (44.8%) | 0.926 |
| Statin, % | 59 (39.6%) | 95 (41.3%) | 0.741 |
| Beta blocker % | 68 (45.6%) | 99 (43%) | 0.619 |
| ACE inhibitor % | 86 (57.7%) | 116 (50.4%) | 0.165 |
| ARB % | 13 (8.7%) | 28 (12.2%) | 0.291 |
| ARNI % | 0 (0%) | 2 (0.9%) | 0.254 |
| Loop diuretic % | 89 (59.7%) | 116 (50.4%) | 0.076 |
| Mineralocorticoid receptor antagonist % | 32 (21.5%) | 41 (17.8%) | 0.379 |
| Hydralazine % | 22 (14.8%) | 29 (12.6%) | 0.548 |
| Imdur % | 23 (15.4%) | 36 (15.7%) | 0.955 |
| Digoxin % | 2 (1.3%) | 4 (1.7%) | 0.762 |

*\* Laboratory data was limited by availability in the hospital medical records and values were not collected for all participants. Data for creatinine and potassium were unreliable on manual adjudication and therefore were not reported.*

**Figure 1a.**
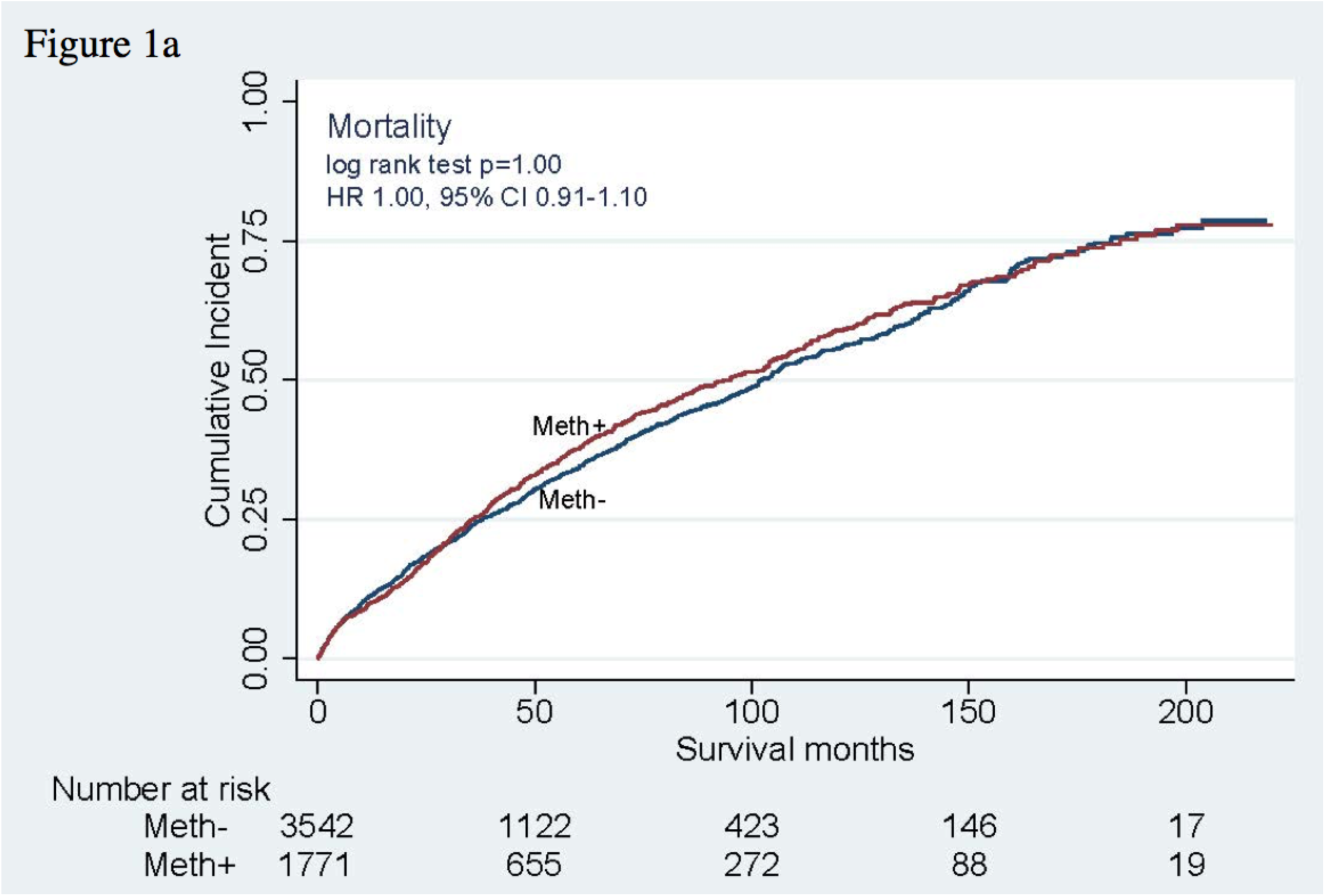
Kaplan-Meier curve demonstrating the cumulative incidence of mortality among the whole cohort stratified by methamphetamine use status with median follow up of 31.6 months (IQR: 9.5 to 69.6 months). HR is adjusted for age and sex.

**Figure 1b.**
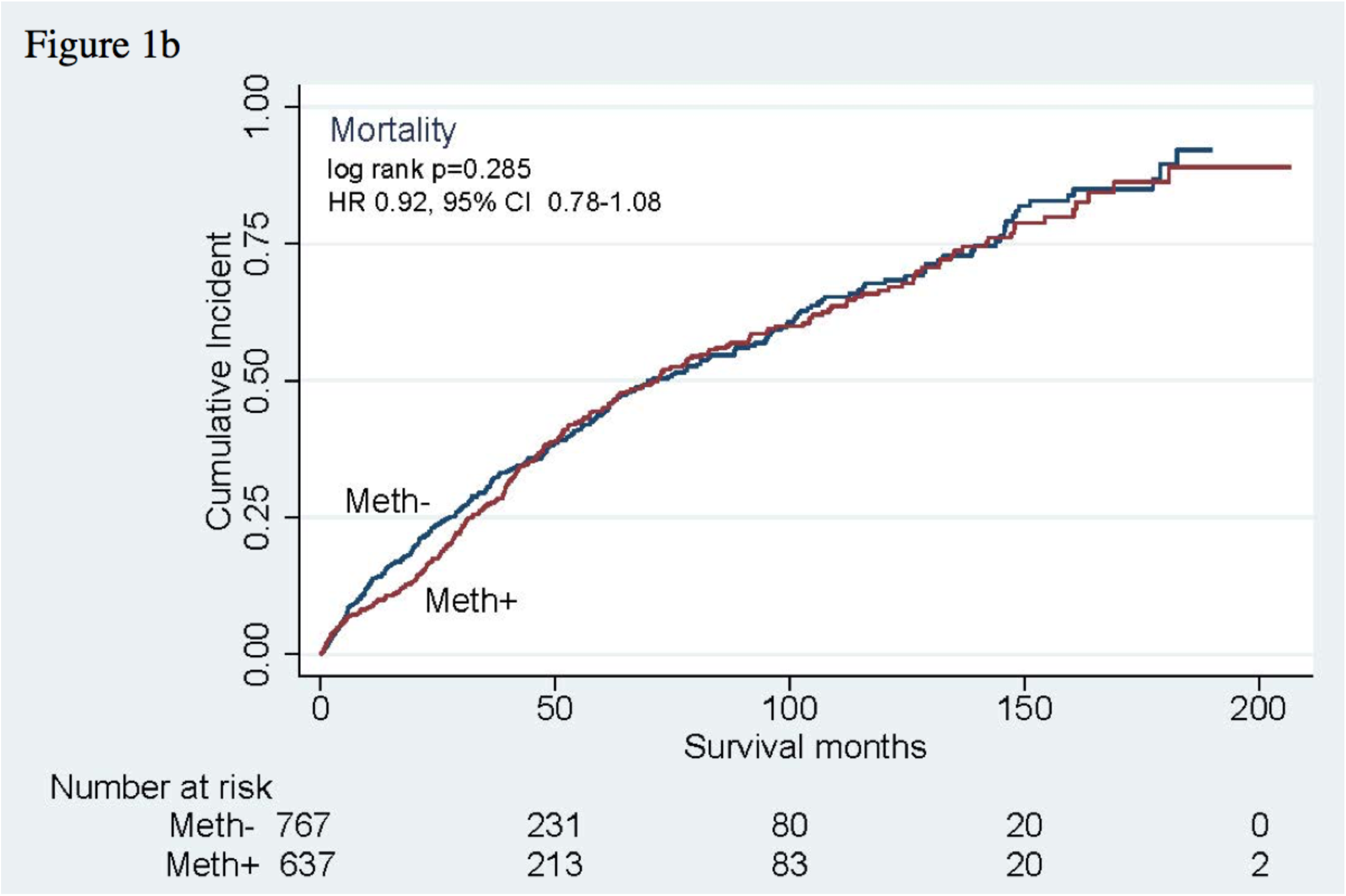
Kaplan-Meier curve demonstrating the cumulative incidence of mortality among the subset of individuals with index HF hospitalization stratified by methamphetamine use status with median follow up of 31.6 months (IQR: 9.5 to 69.6 months). HR is adjusted for age and sex.

After adjusting for demographic characteristics and use of other substances that may confound the effect of methamphetamine use on heart failure outcomes, methamphetamine use was associated with lower mortality compared to individuals with heart failure due to other causes by the end of 1 year and 5 years **(eTable 1).** As a sensitivity analysis, a stepwise backward selection approach was used to consider adjusting for other comorbid conditions; that model included age, race, gender, coronary artery disease, hepatitis B virus, chronic kidney disease, and history of sustained ventricular tachycardia and resulted in similar effect estimates for mortality (**eTable 1).** We did a subgroup analysis of mortality by type of HF and found similar results (HFrEF: HR 0.81, 95% CI 0.67, 0.97, p=0.02; HFpEF: HR 0.74, 95% CI 0.62, 0.89, p<0.01) suggesting that type of heart failure did not significantly influence the association between methamphetamine use and mortality.

In a bivariable analysis of clinical conditions associated with mortality among individuals with and without methamphetamine use, the presence of an outpatient visit within 30 days of hospital discharge was associated with lower mortality among both individuals with history of methamphetamine use (HR 0.77; 95% CI 0.65, 0.92; p=0.004) and without history of methamphetamine use (HR 0.55; 95% CI 0.48, 0.63; p<0.0001).

### Hospital readmissions by methamphetamine use status

Among individuals with an index HF hospitalization, those with methamphetamine use had significantly higher risk of heart failure readmission (52.0%; n=324 vs 47.3%; n=336, HR 1.91, 95% CI 1.33, 2.75 p<.0001) and all cause readmission (57.4%; n=357 vs 50.6%; n=359, HR 1.55, 95% CI 1.13, 1.88, p=0.003) over a median follow up time of 30.8 months (IQR: 11.3 to 63.9 months) as demonstrated in **Figures 2a and 2b**. Results of 30-day, 90-day, and 1 year readmissions are summarized in **Table 3**. Individuals with methamphetamine use had consistently higher heart failure readmission rate at 30 days (RR 1.92, 95% CI 1.36-2.70, p<0.001), 90 days (RR 1.69, 95% CI 1.35-2.12, p<0.001), and 1 year (RR 1.61, 95% CI 1.36-1.91, p<0.001). Methamphetamine use was also associated with increased all-cause readmission rate at 30 days (RR 1.46, 95% CI 1.16-1.83, p<0.001), 90 days (RR 1.41, 95% CI 1.21-1,64, p<0.001), and 1 year (RR 1.29, 95% CI 1.16-1.43, p<0.001).

**Figure 2a.**
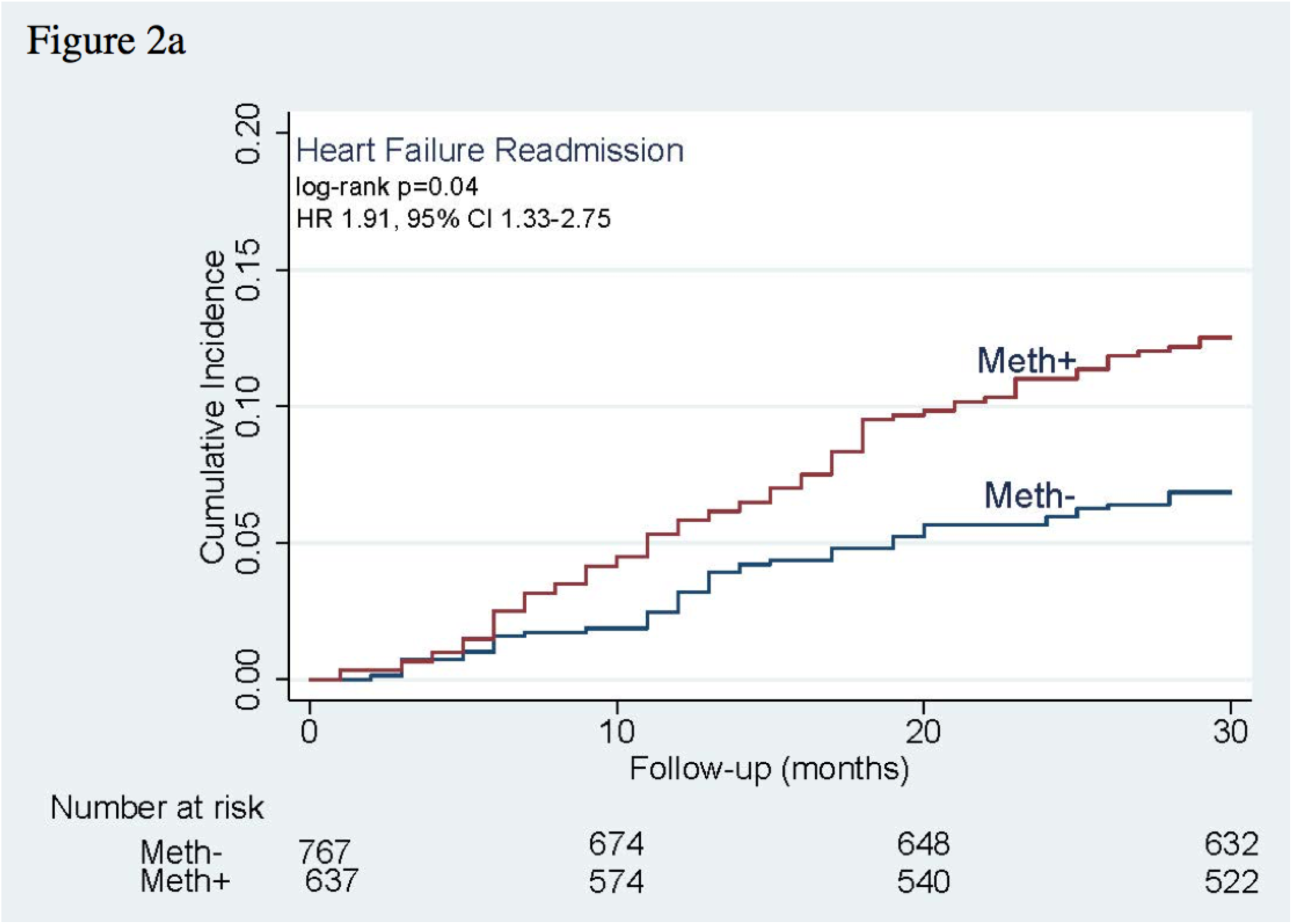
Kaplan-Meier curve demonstrating the cumulative incidence of heart failure readmission among the subset of individuals with an index HF hospitalization stratified by methamphetamine use status with median follow up of 30.8 months (IQR: 11.3 to 63.9 months). HR is adjusted for age and sex.

**Figure 2b.**
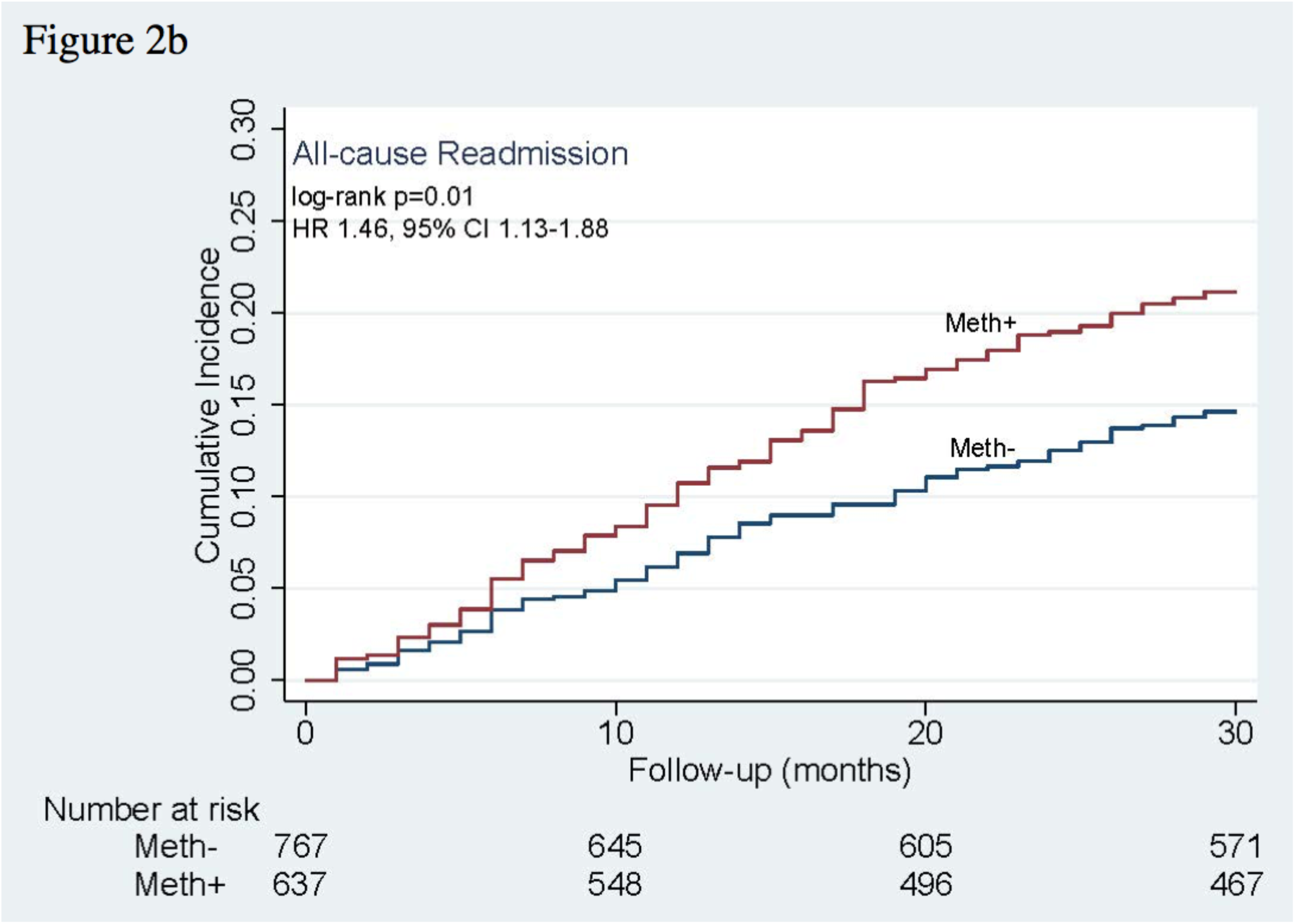
Kaplan-Meier curve demonstrating the cumulative incidence of all cause readmission among the subset of individuals with an index HF hospitalization stratified by methamphetamine use status with median follow up of 30.8 months (IQR: 11.3 to 63.9 months). HR is adjusted for age and sex.

**Table 3.**
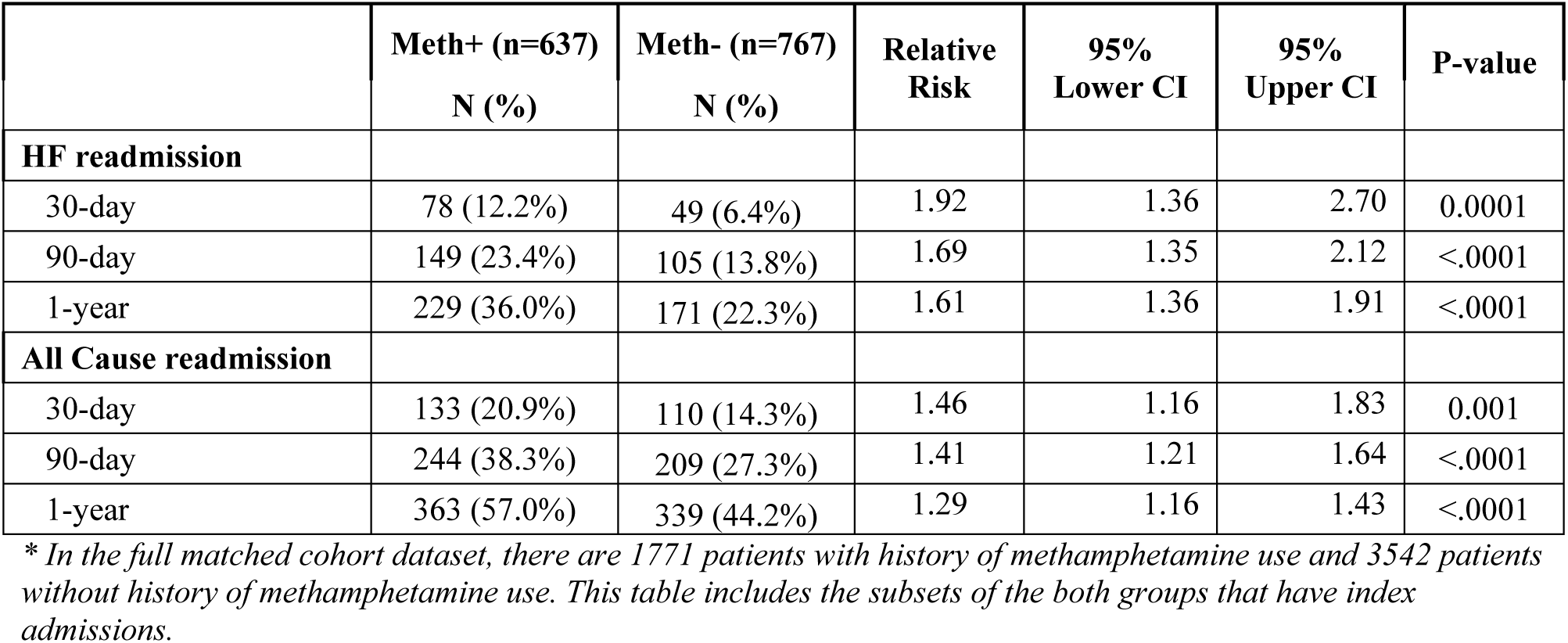
Primary readmission outcomes stratified by methamphetamine use status over study period.

After adjustment for demographic characteristics and use of other substances, methamphetamine use was associated with higher heart failure readmission hazard compared to individuals with heart failure due to other causes (adjusted HR 1.55, 95% CI 1.03, 2.33, p=0.035) but not significantly higher all cause readmission hazard (adjusted HR 1.19, 95% CI 0.89, 1.58, p=0.233) (**eTable 1)**. We did a stratified analysis of mortality by type of HF and again found similar results for HF readmissions (HFrEF: HR 1.63, 95% CI 1.01, 2.65, p=0.05; HFpEF: HR 0.88, 95% CI 0.33, 2.38, p=0.80) and all cause readmissions (HFrEF: HR 1.05, 95% CI 0.74, 1.49, p=0.80; HFpEF: HR 1.59, 95% CI 0.84, 2.99, p=0.15) suggesting that type of heart failure did not significantly influence the association between methamphetamine use and readmission.

Hospitalization characteristics of individuals with an index HF hospitalization and subsequent hospital readmission are summarized in **eTable 2.** Individuals with methamphetamine use were significantly more likely to require ICU level care during the hospitalization or to leave the hospital against medical advice.

## Discussion

Among a large cohort of individuals with heart failure within a municipal health system over a 19-year period, there was no difference in mortality between individuals with and without methamphetamine use. However, methamphetamine use was associated with significantly higher risk of both HF and all-cause readmissions over median follow up of 3 years. This study adds to a small body of literature on this important topic.^5,6^

### Possible Explanations for the Methamphetamine Mortality Paradox

While there was no overall difference between groups in mortality, in adjusted analyses, individuals with heart failure and methamphetamine use had significantly lower mortality as compared to individuals with heart failure due to other causes. This apparent methamphetamine-mortality “paradox” is an interesting finding and is consistent with the findings reported by Thomas et al.^6^ Several hypotheses may explain this unexpected finding. In this study, methamphetamine use was defined as “ever use,” rather than based on dosage exposure over time, due to EHR data limitations. However, methamphetamine use is likely a modifiable risk factor with a percentage of the population stopping use over the follow up period. Cessation of substance use exposure may improve cardiac function and is also associated with reduced mortality, as demonstrated Zhao et al.^7^ Interestingly, in an exploratory analysis we also found that presence of hypertension and obstructive sleep apnea appeared to be protective against mortality. It is possible that the presence of hypertension resulted in greater ability to initiate and uptitrate guideline-directed medical therapy for heart failure, and the presence of obstructive sleep apnea signaled engagement with care to establish this diagnosis.

Furthermore, individuals with heart failure and methamphetamine use had significantly increased risk for short-term and long-term readmissions for heart failure and all causes. Increased contact with the medical system due to higher rates of follow up within 30 days and more frequent readmissions may have resulted in improved heart failure education and medication optimization. Indeed, individuals with methamphetamine use had higher odds of being prescribed guideline-directed medical therapy for heart failure and cardiovascular medications including aspirin and statin compared to individuals without history of methamphetamine use. Higher use of GDMT may explain this paradox, although we did not conduct formal mediation analysis to ascertain the effects independent of GDMT use due to high likelihood of residual confounding.

### Prior Studies of Methamphetamine-Associated HF

Our study builds on the prior work by Thomas et al who found that methamphetamine use was associated with higher rates of heart failure readmissions but a lower 10-year mortality rate in individuals with heart failure and methamphetamine use compared to individuals in the control population.^6^ However, they obtained mortality data solely from the electronic health record, which is likely to be biased by differential misclassification of the primary outcome as individuals with methamphetamine use may be more likely to die outside of the hospital. Additionally, the cohort was not age-matched which could explain the results as the cohort of individuals with methamphetamine-associated heart failure was significantly younger than the control population. A second study by the same group used data from the National Inpatient Sample to demonstrate that HF hospitalizations associated with methamphetamine use are increasing over the past two decades, especially compared to HF hospitalizations associated with use of other substances like cocaine or alcohol.^5^ However, the National Inpatient Sample lacks data on the type of heart failure (reduced vs preserved ejection fraction), pertinent cardiovascular comorbidities, or outcomes like readmission or mortality—characteristics that are vital to understand and treat patients at highest risk of poor outcomes.

Finally, Zhao et al compared the clinical characteristics and outcomes in individuals with reversible versus persistent methamphetamine-associated cardiomyopathy based on change in LVEF over time. They found that individuals with reversible methamphetamine-associated cardiomyopathy had significantly higher cumulative survival compared to individuals with persistent methamphetamine-associated cardiomyopathy.^7^ However, this was a small study of only 357 individuals in which all subjects had exposure to methamphetamine use. Given the well-known contribution of heart failure to readmission and mortality, we chose to study the variable impact of methamphetamine exposure in a population of individuals with a diagnosis of heart failure.

Compared to the other studies published in the literature, our study includes a larger, more diverse cohort over a long follow-up period. We also present key heart failure characteristics including type of heart failure and use of GDMT in order to delineate the role of methamphetamine use in driving clinical heart failure outcomes, significantly adding to our current understanding of this disease. Additionally, we linked our data to national death records for ascertainment of mortality outside of this health system.

### Methamphetamine users more likely to have HFrEF

We found that individuals with heart failure and methamphetamine use were significantly more likely to have reduced ejection fraction as compared to preserved ejection fraction consistent with prior reports in the literature.^8^ In a study of 21 individuals with history of methamphetamine use by Wijetunga et al, 84% underwent echocardiography and had findings of dilated cardiomyopathy and global ventricular dysfunction.^9^ In another small study by Reddy et al of 140 individuals with heart failure, in which 50% had with methamphetamine-associated cardiomyopathy, methamphetamine use was associated with significantly lower LV ejection fraction.^8^ However, it is important to note that all patients in that study had reduced ejection fraction. Further examination of echocardiographic and histologic parameters of heart failure in a large population of individuals with history of methamphetamine use and heart failure will further elucidate the mechanism of methamphetamine-associated cardiomyopathy.

However, in a stratified analysis, we found that the type of heart failure (HFrEF vs HFpEF) did not significantly influence the association between methamphetamine use and mortality or hospital readmission. Differences in heart failure outcomes by EF (regardless of methamphetamine use status) have been reported in the literature. A study by Shah et al of 39,982 patients admitted for heart failure at 254 hospitals found that individuals with HFrEF and heart failure with borderline EF had higher cardiovascular and HF readmission rates as compared to individuals with HFpEF, but no difference in mortality among the groups.^10^ It is important to note, however, that this study did not characterize methamphetamine use patterns among individuals included in the study. This is the first study to our knowledge that describes clinical heart failure outcomes of individuals with and without history of methamphetamine use in which ejection fraction is reported. Given major differences in guideline-directed medical therapy by heart failure type, our paper adds key information to the existing literature about the role of HF type and methamphetamine use on clinical HF outcomes.

### Methamphetamine and Other Cardiotoxic Substance Use

Among individuals with history of methamphetamine use, concomitant use of other substances including cannabis, cocaine, and alcohol significantly increased the risk of heart failure and all cause readmission. This is not surprising, as each individual substance has been shown to have cardiotoxic effects including the development of arrhythmias, heart failure, and myocardial infarction.^11–13^

### Coronary Artery Disease Among Individuals with Methamphetamine Use and Heart Failure

Coronary artery disease significantly increased risk of heart failure and all cause readmissions among individuals with history of methamphetamine use, but not among individuals without history of methamphetamine use. This may be explained by methamphetamine’s role as a sympathomimetic agent resulting in increased myocardial oxygen demand; in the setting of pre existing coronary artery disease, this additional stressor may result in chronic ischemia and increased predisposition for heart failure decompensation.^8^

### Insurance and Follow Up Care and Outcomes

Predictors of mortality were similar between both groups. It is important to note that insurance status—particularly individuals who were uninsured or insured by Medicaid—had significantly higher likelihood of mortality over the 5 year follow up period. This highlights the implications of socioeconomic status on heart failure mortality, and underscores the need for public health interventions to support vulnerable populations living with a diagnosis of heart failure to improve outcomes. Further, presence of a follow-up visit within 30 days of hospital discharge was associated with lower mortality for both individuals with and without history of methamphetamine use. In a prior study of the association between follow up after heart failure hospitalization and risk of readmission in the Kaiser Permanente Northern California health system, post-discharge follow up within 7 days was associated with lower odds of readmission, but later outpatient follow up between 8 and 30 days did not significantly change odds of readmission.^14^ This study did not examine the impact of post-discharge follow up on mortality. It is interesting that post-discharge follow up within 30 days in our study did not show differences in heart failure or all cause readmissions, but did significantly decrease likelihood of mortality. This may be explained by differences in timing (7 vs 30 days), or in socioeconomic status between the safety-net patient populations in our study as compared to the Kaiser Permanente population. Public policy efforts aimed at improving access to follow up care may improve survival and should be pursued at the local and national level.

### Limitations

The results of this study should be interpreted considering certain limitations. First, our data only represent the patient population at a single municipal health system with a robust public insurance system and may not be generalizable to the broader community population. Further study of the impact of methamphetamine use on heart failure outcomes in other populations is necessary given the widespread prevalence of methamphetamine use. Second, this study did not specifically exclude those with methamphetamine use with other possible etiologies for heart failure, but rather considered the total effect of methamphetamine use among individuals with heart failure. We accounted for this with multivariable analysis of comorbid conditions to adjust for their impact on outcomes. Third, all data was queried from the electronic health record (EHR) using ICD and CPT diagnosis and procedure codes, which are inherently imperfect. To counteract this, a single physician manually adjudicated a random sample of the data for accuracy. Furthermore, this study only included those with methamphetamine use who were diagnosed with clinical heart failure, and did not compare outcomes to a control group with methamphetamine use without heart failure or the general population without heart failure or methamphetamine use. Additionally, prior studies have demonstrated that cessation of methamphetamine use can result in reversal of methamphetamine-associated cardiomyopathy if cessation precedes severe cardiac chamber dilatation.^7^ However, we were unable to characterize duration of methamphetamine use in this large cohort due to availability of data. There was also a high degree of loss of follow-up, which was greater among those without methamphetamine use. Finally, the readmission outcomes are limited to the municipal health system, and it is possible that readmissions to other health systems could be different by methamphetamine use status.

### Conclusion

Among individuals with heart failure and methamphetamine use, we found that there was significantly higher risk of hospital readmission for heart failure and all causes, but no significant difference in mortality. There was no significant difference in these outcomes in a stratified analysis by HF subtype. This study is the first to our knowledge to describe clinical characteristics of individuals with methamphetamine use and heart failure, and to comprehensively report on clinical outcomes including mortality and hospital readmission over a prolonged follow up period.

Given the rapidly rising prevalence of methamphetamine use, and the known cardiovascular complications resulting from stimulant use, it is important to understand clinical characteristics of this population and the associated heart failure outcomes related to methamphetamine use. Understanding the clinical characteristics that predict worse outcomes in this population can also help to guide future public policy efforts at the local, state, and national level. Further study of the mechanisms by which methamphetamine results in cardiomyopathy can guide novel therapeutic strategies to treat this vulnerable population.

## Data Availability

The data used in the analyses of this manuscript can be made available upon reasonable request.

## Notes

### Competing Interest Statement

The authors have declared no competing interest.

### Funding Statement

No external funding was received to support this study.

### Author Declarations

The University of California San Francisco institutional review board approved this study.

